# Deep Covid - Covid Diagnosis Using Deep Neural Networks and Transfer Learning

**DOI:** 10.1101/2021.05.20.21257387

**Authors:** Abhinav Sagar

**Author notes:** Website of author - https://abhinavsagar.github.io/.

## Abstract

Coronavirus is a global emergency as of May 2021. If not acted upon by drugs at the right time, coronavirus may result in the death of individuals. Hence early diagnosis is very important along the progress of the disease. This paper focuses on coronavirus detection using x-ray images, for automating the diagnosis pipeline using convolutional neural networks and transfer learning. This could be deployed in places where radiologists are not easily available in order to detect the disease at very early stages. In this study we propose our deep learning architecture for the classification task, which is trained with modified images, through multiple steps of preprocessing. Our classification method uses convolutional neural networks and transfer learning architecture for classifying the images. Our findings yield an accuracy value of 91.03%, precision of 89.76 %, recall value of 96.67% and F1 score of 93.09%.

## 1 Introduction

The risk of coronavirus is immense for many, especially in developing nations where billions face poverty and rely on polluting forms of energy. In such regions, the problem is further increased due to the lack of medical resources and equipment. For these populations, accurate and fast diagnosis is very important. It can guarantee timely access to treatment and save much needed time and money for those already experiencing poverty. Traditionally classifying and detecting coronavirus has been done by trained radiologists. It is very common for radiologists to make a mistake as to the naked eye there are cases where coronavirus could be missed which is also known as false negatives. In other cases, radiologists could diagnose someone as having coronavirus even if he doesn’t have it. Both of the above cases should be avoided which is where convolutional neural networks can help.

Convolutional neural network models have been designed, and experiments were performed using them by grid searching over the hyperparameter space. This process demands huge computational burden, time and skilled expertise. Hence a novel but simple model is required to automatically perform optimal classification tasks with convolutional neural network architecture. The proposed technique utilizes a set of neurons to convolve a given image and extract relevant features from them. Demonstration of the efficacy of the proposed method with the minimization of the computational cost was conducted and compared with the existing state-of-the-art coronavirus classification networks.

## 2 Related Work

There have been previous studies done regarding coronavirus detection with chest x-rays via machine learning with the use of heat maps, and differentiation of pulmonary pathology. While the mentioned conventional and radiological methods might be effective, our study presents a deep learning approach to this coronavirus classification. Looking at the state of art, there have been two previous similar experimentations on this task.

The initial one uses Long Short Term Memory (LSTM) architectures for finding interdependencies among the X-ray data. While their study focuses on 14 interdependent diseases, our study focuses merely on coronavirus. However, due to their experimentation for extracting 14 different diseases with one model, they have merely been able to reach an accuracy of 71.3%. Furthermore, LSTM uses multiple images for classifying a single image, whereas our proposed experimentation and model only need pre trained neural network weights for classifying images one by one. Additionally, our accuracy upon experimentation yielded 91.03%, which uses the same dataset.

The second experimentation was conducted with similar means to ours. They used a 121 layer convolutional network for feature map acquisition, alongside statistical methods (standard deviation and mean calculation) for image preprocessing. Our preprocessing methods are similar to real life applications, unlike statistical means that might be ineffective when a wide range of data is present. Finally, our proposed architecture yields an accuracy of 91.03%, while their study yielded an accuracy of 76.8%.

Other than the above mentioned papers on coronavirus classification, Chest X-ray images have been widely subjected to experimentations with convolutional neural network architectures, as well as other image classification techniques. Bone structures were segmented within a paper. This paper presents a segmentation method which utilizes additional steps after the classification algorithm.

As a regular Convolutional neural network classifies the image as a whole, such segmentation methods utilize pixelwise classification, which, in the end, applies a deconvolutional layer for classifying each pixel one by one and eventually separating different objects within an image, bones being the most prevalent ones for the mentioned task. In another research aiming to conduct early detection not for coronavirus but for thorax disease through weekly classifications with convolutional neural networks. This paper successfully detects patterns for patients who have thorax disease or one that might have the mentioned disease.

## 3 Proposed Method

### 3.1 Dataset

The dataset is organized into 3 folders for training, validation and testing respectively. It also contains subfolders for each image category i.e. coronavirus and normal images. There are 5,863 X-Ray images and 2 categories coronavirus and normal. Chest X-ray images were selected from retrospective cohorts of pediatric patients of one to five years old from Guangzhou Women and Children’s Medical Center, Guangzhou. Fig 1 and Fig 2 shows the sample normal and coronavirus images respectively present in the dataset. As can be seen, to the naked eye it is very difficult to tell the differences between the two.

**Figure 1:**
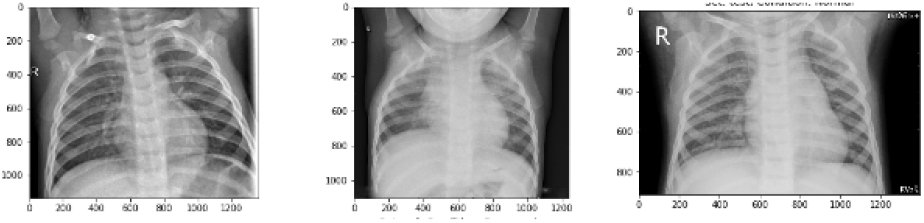
Sample images without covid.

**Figure 2:**
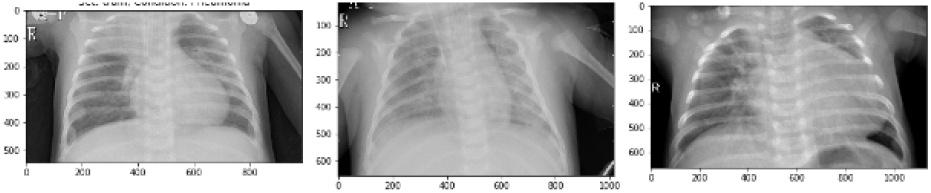
Sample images with covid.

### 3.2 Network Architecture

We split the data-set into three sets — train, validation and test sets. We used data augmentation like rotate, shearing, zooming, horizontal and vertical flipping, contrast, shiftRGB, shiftHSV, grayscale, equalize, resize, multiply and brightness change to increase the dataset size to almost double the original dataset size. This operation helps the model prevent overfitting thus making it more generalizable to unseen images in the test set. The data augmentation parameters used in our work is shown in Table 1.

**Table 1:** Data augmentation parameters

| Transformation | Parameters |
| --- | --- |
| Rotation | -20 degrees to +20 degrees |
| Horizontal Flip | Randomness 50% |
| Vertical Flip | Randomness 50% |
| Horizontal Shift | Upto 12 pixels |
| Vertical Shift | Upto 12 pixels |
| Gaussian Blurring | Upto 2 pixel |

We used position augmentation like scaling, cropping, flipping, padding, rotation, translation, affine transformation. These operations are shown in Fig 3.

**Figure 3:**
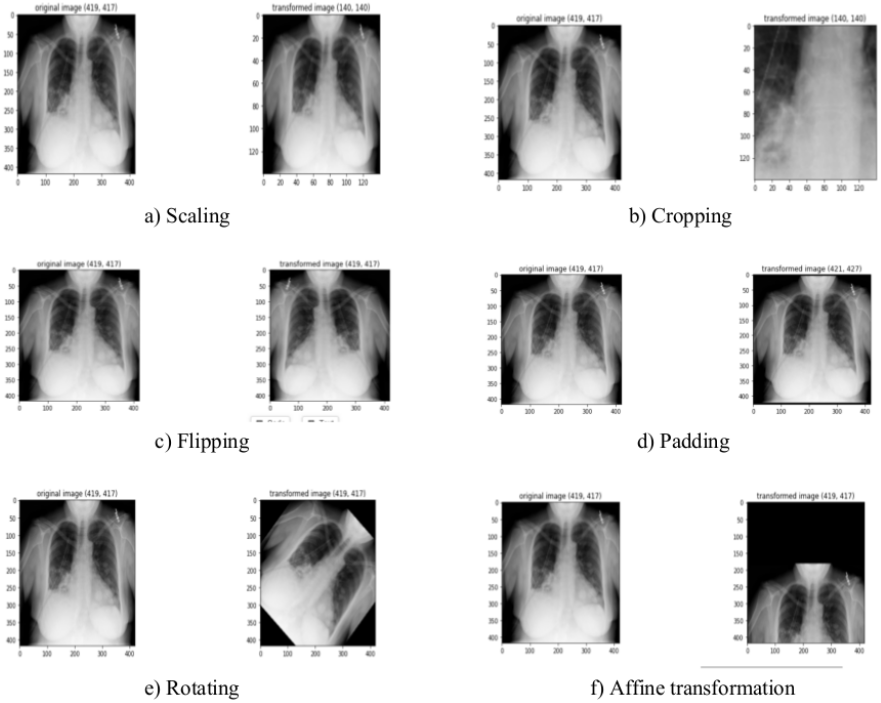
Position augmentation techniques.

Color augmentation techniques like brightness, contrast, saturation and hue are used. These operations are shown in Fig 4.

**Figure 4:**
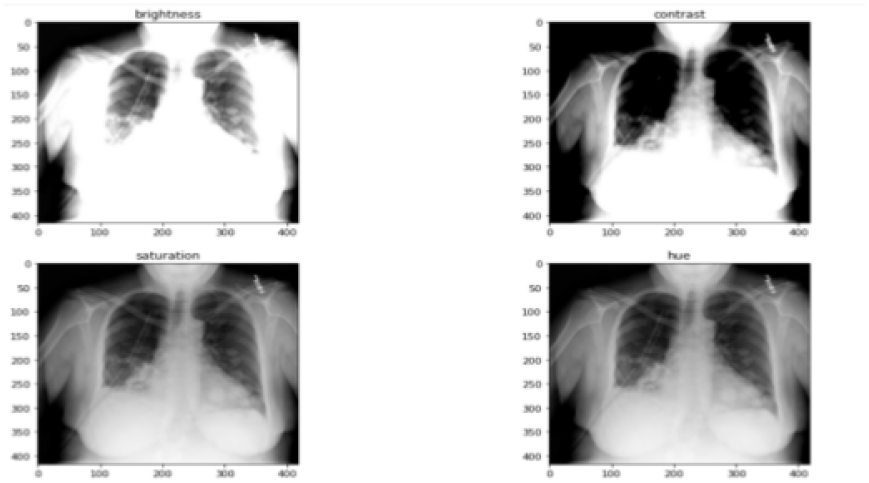
Colour augmentation techniques.

We defined a couple of data generators: one for training data, and the other for validation data. A data generator in this case is used for loading the required amount of data which is often a mini batch of images directly from the source folder, converting them into training data which are fed to the model and training targets denoting the labels.

We also converted the images to HSV (Hue, Saturation and Value) format. This conversion made the neural network avoid overfitting and thus make it more generalizable to unseen results. We compared the model using the original images and found the images converted to HSV to give better results on all metrics including F1, accuracy and ROC. Sample images converted to HSV format are shown in Fig 5.

**Figure 5:**
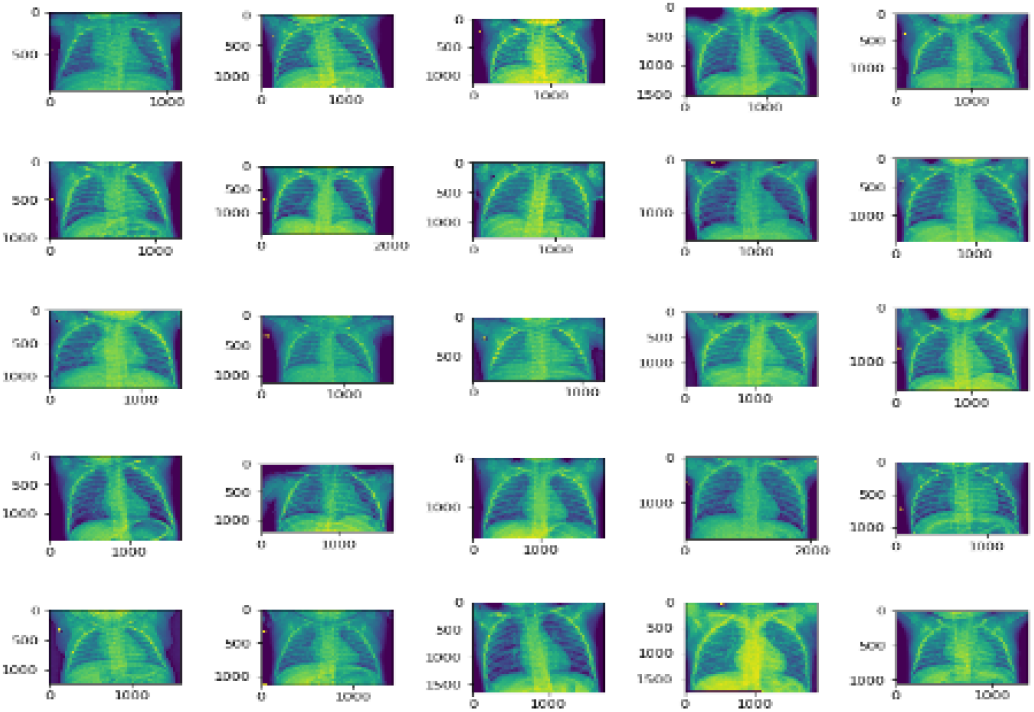
Sample images in HSV format.

Digital images are composed of two-dimensional integer arrays denoted using pixel values that represent individual components of the image. The number of bits used to represent these pixels determines the number of gray levels used to describe each pixel. The pixel values in black-and-white images can be either 0 (black) or 1 (white), representing the darker and brighter areas of the image.

If n bits are used to represent a pixel, then there will be 2n pixel values ranging from 0 to (2n − 1). Here 0 and (2n − 1) correspond to black and white, respectively, and all other intermediate values represent shades of gray. The histogram of the pixel intensities variation is shown in Fig 6. The intensity is higher in the right part, less in the left and varies with a lot of variations in the middle.

**Figure 6:**
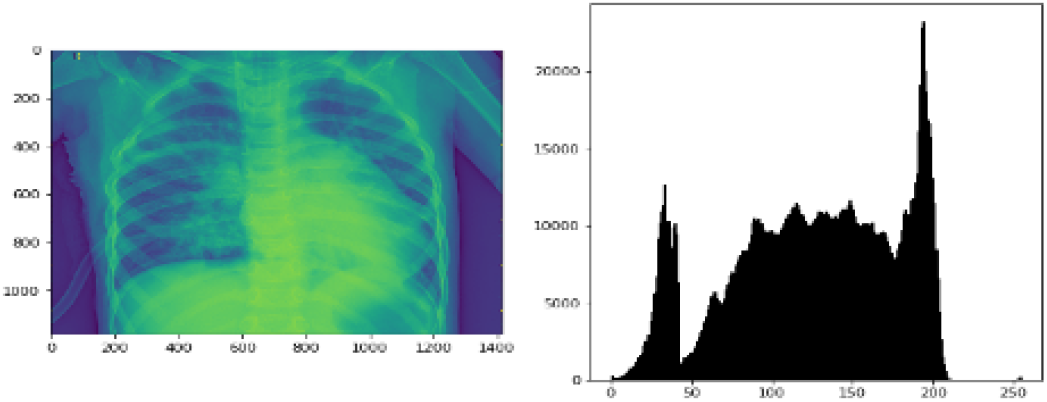
Histogram of Pixel Intensities.

### 3.3 Optimization

The feature maps help in explaining what the model is learning at every layer. As the depth increases, the model is able to learn more spatial information. In other words, the neural networks go on from learning edges and blobs in the first layer to complete objects in the last layers.

Visualization of feature maps is important because this makes hyperparameter tuning easier, since when we see an error made by the neural network then we can know what is making the algorithms going wrong. The functionality and expected behaviour of the networks can be explained, especially to non-technical stakeholders who would often demand explanation before accepting the results.

Also we can further extend and improve the overall design of our models since we’d have knowledge of the current design using the learned filters and we can improve that for the future models. By visualising the learned weights we can get some idea as to how well our network has learned. For example, if we see a lot of zeros then we’ll know we have many dead filters that aren’t doing much for our network which means we need to do some pruning for model compression. The activation of the feature maps for the first three convolutional and max pooling layers is shown in figure 7.

**Figure 7:**
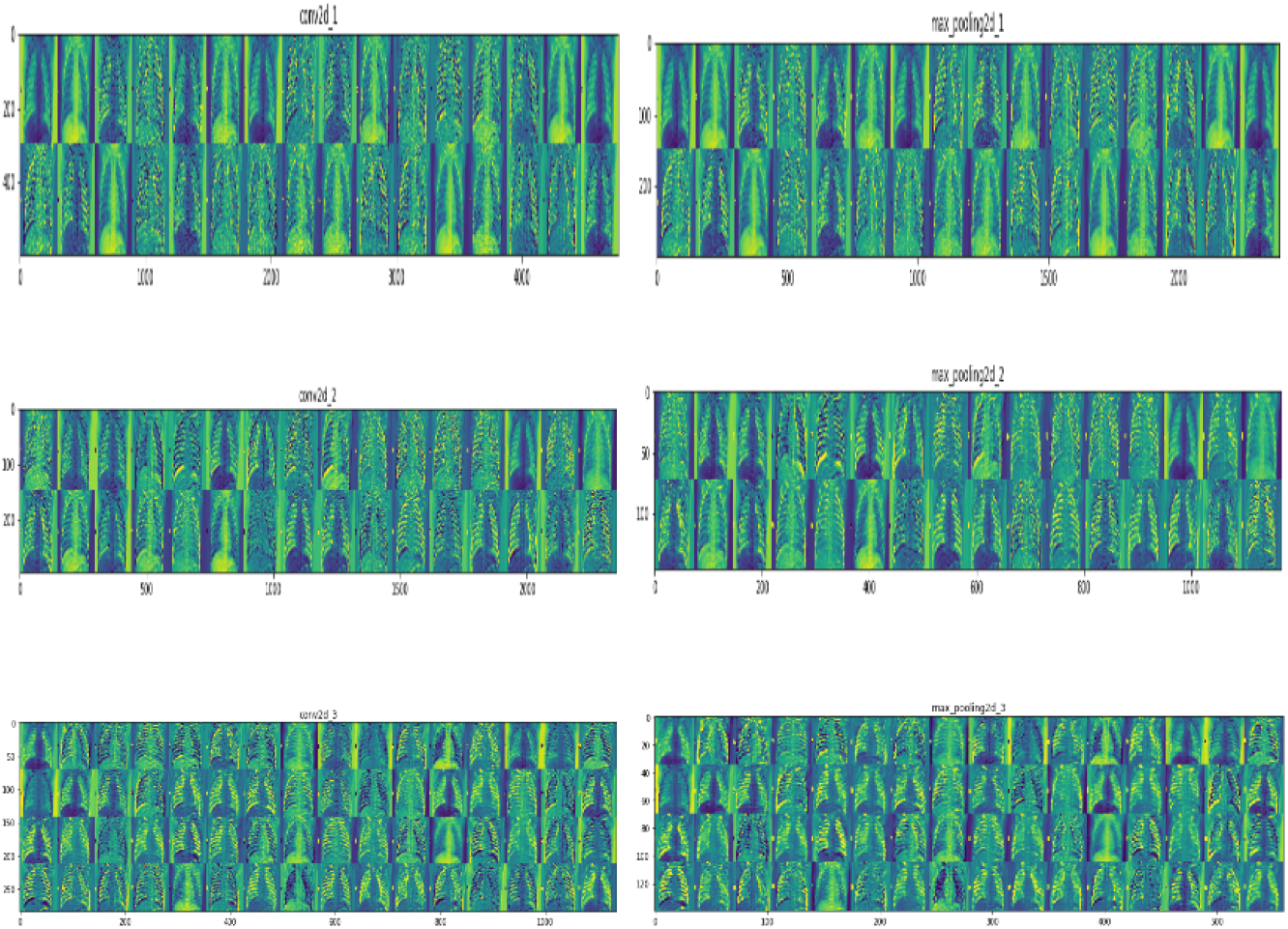
Sample images in HSV format.

In between the convolutional and max pooling layers for each block, we used 30% dropouts to reduce overfitting. Activation function used was Relu throughout except for the last layer where it was Sigmoid as this is a binary classification problem. We used both Adam as the optimizer and binary cross-entropy as the loss function.

We tried with pre trained models like Inception v3, InceptionResNet v2 and ResNet 152 by fine tuning the last layers of the network. We used two dense layers with 64 neurons and 2 neurons respectively. We trained the model for 20 epochs with a batch size value of 16 by doing a grid search for the optimal hyper-parameters values like learning rate, batch size, optimizer and pre-trained weights.

We defined the optimal as the maximum value achieved for F1 score which is the weighted average of precision and recall. Since the F1 value takes both false negative and false positive and penalizes them in a single term, hence it would be the most accurate measure of the classifier. We used model checkpoint while training the neural network and early stopping when the validation loss started increasing. These architectural and training details are presented in the next subsection.

We used ModelCheckpoint. Often many iterations are required, when training requires a lot of time to achieve a good result. In this case, it is better to save a copy of the best performing model only when an epoch that improves the metrics ends.

We also used EarlyStopping. Sometimes, during training we can notice that the generalization gap (i.e. the difference between training and validation error) starts to increase, instead of decreasing. This is a symptom of overfitting that can be solved in many ways. (reducing model capacity, increasing training data, data augmentation, regularization, dropout, etc). Often a practical and efficient solution is to stop training when the generalization gap is getting worse. Fig 28 shows early stopping.

### 3.4 Results

The loss vs epochs and accuracy vs epochs is plotted in Fig 8. The loss has converged on both the training and the validation set in 60 iterations. The accuracy on the training set is 100% and on validation set is 95%. Since the model was not shown the images on the validation set while training, the accuracy is quite good.

**Figure 8:**
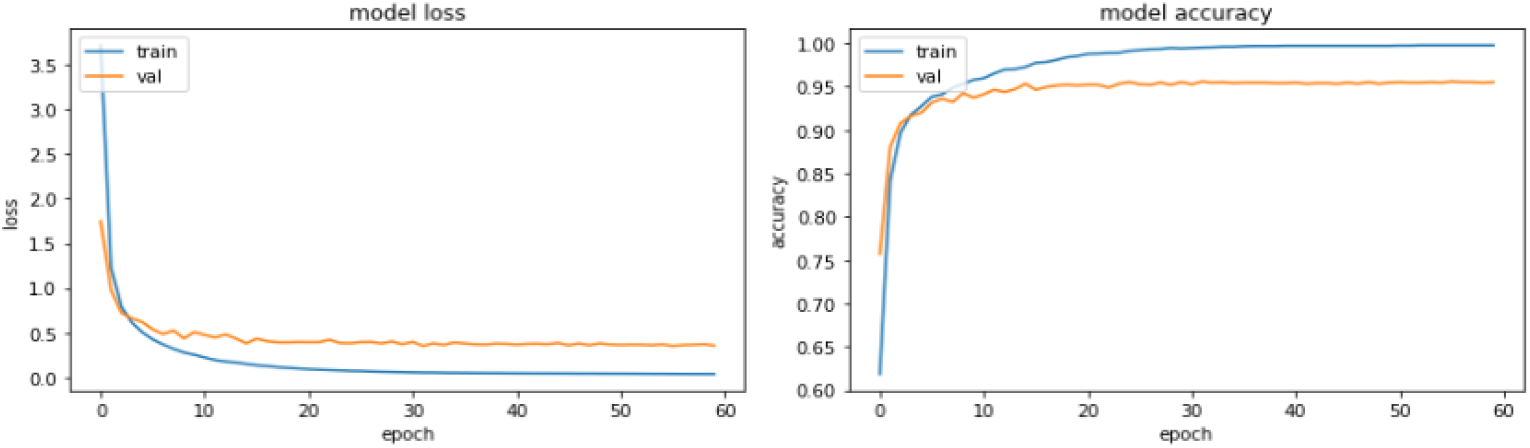
Loss and accuracy vs epochs.

We also used ROC-AUC curves to evaluate the classifier. The 45 degree line is the random line, where the area under the curve or AUC is 0.5. The further the curve from this line, the higher the ROC-AUC and better the model is at classifying. The highest a model can get is an AUC value of 1, where the curve forms a right angled triangle. This denotes that the classifier is perfect. The ROC curve can also help in debugging the model misclassifications. The ROC-AUC curve of the binary classifier is shown in Fig 9:

**Figure 9:**
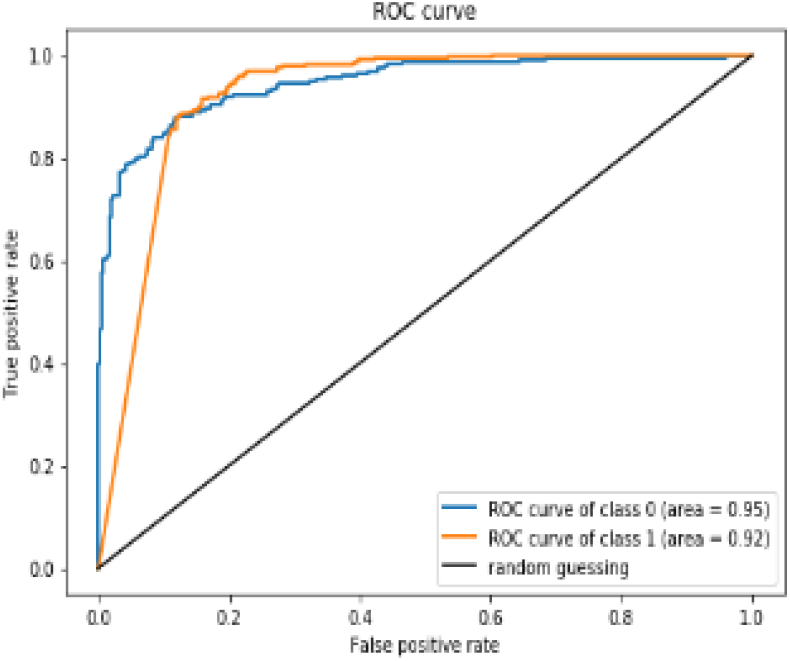
ROC-AUC curve.

The evaluation and results of trained models is calculated by common classification metrics which are defined as follows. The acronyms, TP, TN, FP and FN refer to true positive, true negative, false positive and false negative. A common and widely used quantity is accuracy, which is only a reasonable measure if the different classes in the dataset are approximately equally distributed. It specifies the percentage of objects that have been correctly classified. We present our final results i.e. True Positive, True Negative, False Positive, False Negative Accuracy, Precision, Recall and F1 Score which is the weighted average for InceptionV3, InceptionResNetV2 and ResNet50 individually in Table 2:

**Table 2:** TP, TN, FP, FN, Precision, recall F1 score values

| Metrics | InceptionV3 | InceptionResNetV2 | ResNet50 |
| --- | --- | --- | --- |
| True Positive | 187 | 188 | 191 |
| True Negative | 368 | 370 | 377 |
| False Positive | 49 | 46 | 43 |
| False Negative | 21 | 21 | 13 |
| Accuracy | 90.67 | 90.28 | 91.03 |
| Precision | 90.08 | 89.81 | 89.76 |
| Recall | 96.37 | 97.12 | 96.67 |
| F1 Score | 92.64 | 93.04 | 93.09 |

## 4 Conclusions

In this paper we demonstrated how to classify positive and negative coronavirus images from a collection of X-ray images. We compared various transfer learning architectures like ResNet50, InceptionV3 and InceptionResNetV2 and found ResNet50 to give the best overall results. For evaluation we used metrics including precision, recall, F1 score and ROC-AUC. Finally we made a comparative analysis using various transfer learning architectures, learning rates, batch size and optimizers. By using grid search hyperparameter optimization, the best results were obtained with a learning rate value of 0.0001, batch size value of 16 and ADAM as the optimizer. Due to shortage of radiologists in the world, it is often the case that coronavirus gets unnoticed. Also many times mistakes are made by radiologists themselves, hence it is a good idea to automate the process for better and more efficient diagnosis.

## Data Availability

The details are mentioned in paper.

## Acknowledgments

We would like to thank Nvidia for providing the GPUs.

## References

B. Ghoshal and A. Tucker. Estimating uncertainty and interpretability in deep learning for coronavirus (covid-19) detection. arXiv preprint 2003.10769, 2020.

O. Gozes, M. Frid-Adar, N. Sagie, H. Zhang, W. Ji, and H. Greenspan. Coronavirus detection and analysis on chest ct with deep learning. arXiv preprint 2004.02640, 2020.

K. He, X. Zhang, S. Ren, and J. Sun. Identity mappings in deep residual networks. In European conference on computer vision, pages 630–645. Springer, 2016.

G. Huang, Z. Liu, L. Van Der Maaten, and K. Q. Weinberger. Densely connected convolutional networks. In Proceedings of the IEEE conference on computer vision and pattern recognition, pages 4700–4708, 2017.

D. S. Kermany, M. Goldbaum, W. Cai, C. C. Valentim, H. Liang, S. L. Baxter, A. McKeown, G. Yang, X. Wu, F. Yan, et al. Identifying medical diagnoses and treatable diseases by image-based deep learning. Cell, 172(5):1122–1131, 2018.

D. P. Kingma and J. Ba. Adam: A method for stochastic optimization. arXiv preprint 1412.6980, 2014.

A. Krizhevsky, I. Sutskever, and G. E. Hinton. Imagenet classification with deep convolutional neural networks. Advances in neural information processing systems, 25:1097–1105, 2012.

P. Rajpurkar, J. Irvin, K. Zhu, B. Yang, H. Mehta, T. Duan, D. Ding, A. Bagul, C. Langlotz, K. Shpanskaya, et al. Chexnet: Radiologist-level pneumonia detection on chest x-rays with deep learning. arXiv preprint 1711.05225, 2017.

A. Sagar. Uncertainty quantification using variational inference for biomedical image segmentation. arXiv preprint arXiv:2008.07588, 2020.

A. Sagar and J. Dheeba. Convolutional neural networks for classifying melanoma images. bioRxiv, 2020a.

A. Sagar and J. Dheeba. On using transfer learning for plant disease detection. bioRxiv, 2020b.

R. R. Selvaraju, M. Cogswell, A. Das, R. Vedantam, D. Parikh, and D. Batra. Grad-cam: Visual explanations from deep networks via gradient-based localization. In Proceedings of the IEEE international conference on computer vision, pages 618–626, 2017.

K. Simonyan and A. Zisserman. Very deep convolutional networks for large-scale image recognition. arXiv preprint 1409.1556, 2014.

Y. Song, S. Zheng, L. Li, X. Zhang, X. Zhang, Z. Huang, J. Chen, R. Wang, H. Zhao, Y. Zha, et al. Deep learning enables accurate diagnosis of novel coronavirus (covid-19) with ct images. IEEE/ACM Transactions on Computational Biology and Bioinformatics, 2021.

N. Srivastava, G. Hinton, A. Krizhevsky, I. Sutskever, and R. Salakhutdinov. Dropout: a simple way to prevent neural networks from overfitting. The journal of machine learning research, 15(1): 1929–1958, 2014.

C. Szegedy, W. Liu, Y. Jia, P. Sermanet, S. Reed, D. Anguelov, D. Erhan, V. Vanhoucke, and A. Rabinovich. Going deeper with convolutions. In Proceedings of the IEEE conference on computer vision and pattern recognition, pages 1–9, 2015.

C. Szegedy, V. Vanhoucke, S. Ioffe, J. Shlens, and Z. Wojna. Rethinking the inception architecture for computer vision. In Proceedings of the IEEE conference on computer vision and pattern recognition, pages 2818–2826, 2016.

C. Szegedy, S. Ioffe, V. Vanhoucke, and A. Alemi. Inception-v4, inception-resnet and the impact of residual connections on learning. In Proceedings of the AAAI Conference on Artificial Intelligence, volume 31, 2017.

A. Veit, M. Wilber, and S. Belongie. Residual networks behave like ensembles of relatively shallow networks. arXiv preprint 1605.06431, 2016.

L. Xu, J. S. Ren, C. Liu, and J. Jia. Deep convolutional neural network for image deconvolution. Advances in neural information processing systems, 27:1790–1798, 2014.

X. Xu, X. Jiang, C. Ma, P. Du, X. Li, S. Lv, L. Yu, Q. Ni, Y. Chen, J. Su, et al. A deep learning system to screen novel coronavirus disease 2019 pneumonia. Engineering, 6(10):1122–1129, 2020.

J. Yosinski, J. Clune, Y. Bengio, and H. Lipson. How transferable are features in deep neural networks? arXiv preprint 1411.1792, 2014.

M. D. Zeiler and R. Fergus. Visualizing and understanding convolutional networks. In European conference on computer vision, pages 818–833. Springer, 2014.

